# Risk factors for outbreaks of COVID-19 in care homes following hospital discharge: a national cohort analysis

**DOI:** 10.1101/2020.08.24.20168955

**Authors:** Chris Emmerson, James P Adamson, Drew Turner, Mike B Gravenor, Jane Salmon, Simon Cottrell, Victoria Middleton, Buffy Thomas, Brendan W Mason, Chris J Williams

## Abstract

**Background:** Adult residential and nursing care homes are settings in which older and often vulnerable people live in close proximity. This population experiences a higher proportion of respiratory and gastrointestinal illnesses than the general population and has been shown to have a high morbidity and mortality in relation to COVID-19.

**Methods:** We examined 3,115 hospital discharges to 1,068 Welsh adult care homes and the subsequent outbreaks of COVID-19 occurring over an 18 week period between 22 February and 27 June 2020. A Cox proportional hazards regression model was used to assess the impact of time-dependent exposure to hospital discharge on the incidence of the first known outbreak, over a window of 7-21 days after discharge, and adjusted for care home characteristics, including size, type of provision and health board.

**Results:** A total of 330 homes experienced an outbreak of COVID-19, and 544 homes received a discharge from hospital over the study period. The exposure to discharge from hospital was not associated with a significant increase in the risk of a new outbreak (hazard ratio 1·15, 95% CI 0·89, 1·47, p = 0·29) after adjusting for care home characteristics. Care home size was by far the most significant predictor. Hazard ratios (95% CI) in comparison to homes of <10 residents were: 3·40 (1·99, 5·80) for 10-24 residents; 8·25 (4·93, 13·81) for 25-49 residents; and 17·35 (9·65, 31·19) for homes of 50+ residents. When stratified for care home size, the outbreak rates were similar for periods when homes were exposed to a hospital discharge, in comparison to periods when homes were unexposed.

**Conclusion:** Our analyses showed that large homes were at considerably greater risk of outbreaks throughout the epidemic, and after adjusting for care home size, a discharge from hospital was not associated with a significant increase in risk.

**Research in context:** *What is already known on this subject:*

- Care home populations experience more respiratory outbreaks than the general population^1^ and older people have been more severely affected by COVID-19, with a case fatality proportion of 2·3% overall but 8% in those aged 70-79 and 14·8% in those aged over 80^2^
- Evidence and modelling suggested that up to half of all COVID-19 fatalities could come from the care home population^3^ and that testing prior to hospital discharge was not always available or undertaken^9^
- Type and use of PPE^6^ and the number of staff employed can have an impact on care home outbreaks of COVID-19^6,7^

*What this study adds:*

- Our analysis found no effect of hospital discharges on care home outbreaks once care home size had been adjusted for. In line with previous studies, larger care homes were much more likely to experience an outbreak

## INTRODUCTION

Care homes are settings in which resident populations typically live in close proximity. Annually, they experience outbreaks of gastrointestinal and respiratory illnesses, including norovirus and influenza, with associated morbidity and mortality; 70% of acute respiratory infection outbreaks in the UK occurred in care homes in the winter of 2018/19^1^. Outbreak-associated infections may be introduced via human sources such as new admissions from home or hospital, via staff or via visitors.

Early evidence from the COVID-19 pandemic, later further corroborated, was that older people were more severely affected, with a case fatality proportion of 2·3% overall but 8% in those aged 70-79 and 14·8% in those aged over 80^2^. An assessment of international evidence from April estimated that in Italy and Spain, over half of reported deaths were in care home residents^3^.

Preliminary studies from April in England found extensive spread among staff and residents in homes reporting incidents, and wide variation in symptom profiles^4^. A Scientific Pandemic Influenza Group on Modelling (SPI-M) paper predicted that nearly all care homes would become affected if current conditions persisted, and indicated a role for staff in introducing infections, particularly where staff worked across more than one home^5^. Recent studies indicate that Personal Protective Equipment (PPE) and number of staff employed^6,7^ have an impact on the number of COVID-19 infections. More recent data from the Care Quality Commission suggest that, to mid-June, 36% of care homes experienced an outbreak (defined as a single laboratory confirmed case)^7^ with a study of care homes across a large Scottish Health Board reporting a figure of 37%^7^.

Early estimates of the impact of COVID-19 in the UK suggested that inpatient and critical care bed capacity could be overwhelmed^8^. Hospitals in the UK prepared rapidly for the increase in cases, including cancellation of elective procedures and expediting discharges to home or social care facilities. Testing for residents scheduled for discharge was not always available or done^9^. Media reports have implicated these discharges as the cause for many of the subsequent outbreaks in care homes^10-13^ but we were unable to locate any studies either published or in preprint that linked data on discharges to outbreaks. Expert commentary on existing data has identified care and non-care staff, visitors and resident discharges from hospital as possible vectors for the introduction of COVID-19 into care homes, particularly where testing is not available,^14^ and the discharge back to their care home of untested SARS-CoV-2 positive individuals has been suggested as a risk factor for outbreaks in these settings^15,16^. Studies reporting evidence from testing all staff and residents in specific care homes has suggested high proportions of asymptomatic cases, particularly amongst older residents, than were initially assumed^7,14,17^. This suggests the risk of importing COVID-19 into care homes via hospital discharge of untested asymptomatic residents has been underestimated.

Several studies have provided further evidence of factors that may have increased the risk of outbreak in care homes. A large survey carried out by the Office for National Statistics suggested frequent use of agency staff or carers and staff working conditions, including provision of sick pay, influence the risk of an outbreak^18^. Two studies have used routine data to consider a range of risk factors, including resident need, evidenced by services provided (e.g. nursing care, dementia care), corporate ownership and pre-COVID-19 outbreak history^7, 19^

Wales had its first case of COVID-19 confirmed on 28 February 2020, and also saw a subsequent rise in cases and outbreaks in care homes. Public Health Wales’s Communicable Disease Surveillance Centre has been undertaking surveillance for outbreaks, most of which have been in care settings, since 2015.

We aimed to use our surveillance framework to test whether the risk of a COVID-19 outbreak in the period following a discharge from hospital to a care home was increased compared to other periods, in order to better understand the sources of infection and prevent further incidents.

## METHODS

The study population was all adults living in residential or nursing care homes in Wales, which has seven health boards and a population of 3,152,879 ^20^ The maximum capacity of all Welsh adult care homes during the study period was 25,661 places. All adult residential and nursing homes in Wales (n=1,073 in Wales during study period) were included in the study and care homes not providing adult care were excluded.

Data on notifications of COVID-19 cases and outbreaks were sourced from Tarian, the all Wales health protection case and incident management system. This patient level data system includes laboratory test data from all laboratories in Wales for notifiable disease causative agents. Data on hospital discharges were sourced from the Patient Episode Database for Wales (PEDW).

We linked data for Welsh care home outbreaks of COVID-19 (14 March to 27 June 2020) with data for discharges from hospital to adult care homes in Wales (22 February to 20 June 2020). Our outcome was the time (from 22 February) to the first laboratory confirmed case of COVID-19 in each care home. We defined a baseline exposure period following a discharge from hospital as 7 to 21 days post discharge. Thus, any first case appearing during this window was recorded as being associated with the discharge event. This window was chosen to approximately account for the potentially incubation and infectious period of an asymptomatic or pre-symptomatic (and thus untested) discharged resident and for subsequent incubation period of cases caused by onward transmission in the home. As described below, testing pathways for outbreak identification were only routinely available for symptomatic care home residents during the study period. We considered this baseline scenario the most likely to capture an outbreak if caused by a discharge event, but also considered a sensitivity analysis in which all 1-week, 2-week and 3-week windows between 0 and 31 days post discharge event were analysed.

The first notification of a case of COVID-19 in a care home in Wales was made to Public Health Wales on 15 March 2020 relating to a specimen collected on 14 March 2020; before this date all homes were thus at risk. Once homes had a case of COVID-19 confirmed by laboratory test result, they were excluded from further analysis. This was due to the considerable uncertainty in assigning subsequent cases to a chain of transmission within the home, or to external exposure.

### Care home outbreak ascertainment

We defined a care home as a premises registered with Care Inspectorate Wales (CIW), and recorded as supporting adults. A list of all care homes in Wales was downloaded from the CIW website on 20 May; this contained 1,073 adult care homes. Data were linked by matching addresses on the CIW registration record with addresses recorded on individual hospital discharges from PEDW, and test result records reported on Tarian. Data on care home capacity and nursing care provision were derived from CIW registration records (CIW website). Data on dementia services were provided by CIW based on a 2019 review of registration records.

The testing policy for care home cases changed during the time period for this analysis. Initially, testing was offered for up to the three most recently symptomatic individuals in homes which had not already recorded a confirmed case. This was increased to up to 5 symptomatic individuals from 15 April, and to all symptomatic residents from 24 April. Due to likely under-ascertainment of cases in the earlier part of this period, we defined a COVID-19 outbreak as one resident testing positive for SARS-CoV-2 whilst resident (consistent with Burton et al.^7^), or within 14 days of being resident. All testing was performed by health boards in Wales and all samples processed by NHS laboratories. From 02 May, all hospital patients were required to have a negative COVID-19 test result before being allowed to be discharged back into a care home.

All PCR positive results for SARS-CoV-2 in Welsh residents are uploaded from the laboratory IT system onto the Tarian system. All Tarian cases to 24 June were extracted from the Tarian system on 25 June 2020, with additional results to 27 June extracted on 27 July 2020. Records with postcodes matching those recorded for CIW-registered care homes were identified and addresses manually matched to ensure only those with care home addresses were included. The date on which the specimen was taken was used as the case date in analysis.

### Hospital discharge ascertainment

Data were extracted from PEDW for the period 22 February (21 days prior to the first case notified to Public Health Wales) to 20 June 2020, the most recent date for which data were available. The extract was made on 04/08/20 and all discharges relating to postcodes matching those recorded for CIW-registered care homes were identified and addresses manually matched to ensure only those with care home addresses were included (2,218 discharge records). In addition, to capture events for which a postcode was inaccurate or not recorded, a search was conducted using presence of known care home name in the first line of the address. This identified a further 913 for a total of 3,131 hospital discharge events. Discharge records relating to five care homes across two sites could not be allocated to a specific home. Records relating to them were therefore excluded from analysis, and the final dataset included 3,115 discharges across 1,068 care homes, with a combined maximum capacity of 25,384 residents. Full details on data cleaning are given in supplementary information.

### Statistical Analysis

We used a Cox proportional hazards regression model^21^ to estimate the effect of discharge on the rate at which homes first became affected by COVID-19. Since we defined the (baseline) exposure period as 7-21 days post discharge, we considered the factor ‘hospital discharge’ as a time-dependent covariate in the model. Thus any home could potentially move back and forth between the at-risk or not at-risk categories over time. Additional covariates investigated were obtained from CIW: size of home, services available (nursing, specialist care for dementia or learning disabilities) and region (health board). Hazard ratios were calculated for the unadjusted univariable models and for the mutually adjusted full model. In our sensitivity analysis, we considered the wide range of possible exposure windows (between 0 and 31 days), controlling for the false discovery rate using q values^22^. We also calculated outbreak event rates per 1000 days of exposure to hospital discharge compared to the event rate per 1000 days unexposed, and stratified these by care home size.

This study period timeline is shown in Figure 1. The overview depicts how the 7-21 day risk period follows a discharge date. Each time there was a discharge to a home, a new risk period was added to the model. Depending on timing of discharges to a home, risk periods in that home could be consecutive (scenario A), not occur (scenario B) or be overlapping (scenario C). In the case of overlapping risk periods, these were considered cumulative in our model, extending the overall risk period. As such, our model accounted for the possibility of care homes having none, some or all risk periods overlapping. Our end point was time to first outbreak, hence if an outbreak occurred in a home (scenarios B, C and D), it was censored at that point.

### Patient and public involvement

Due to the nature of this study (analysis of routine data) and the imperative to analyse data and report results rapidly to support public health responses to COVID-19, it was not possible to involve patients and the public in this study.

**Figure 1.**
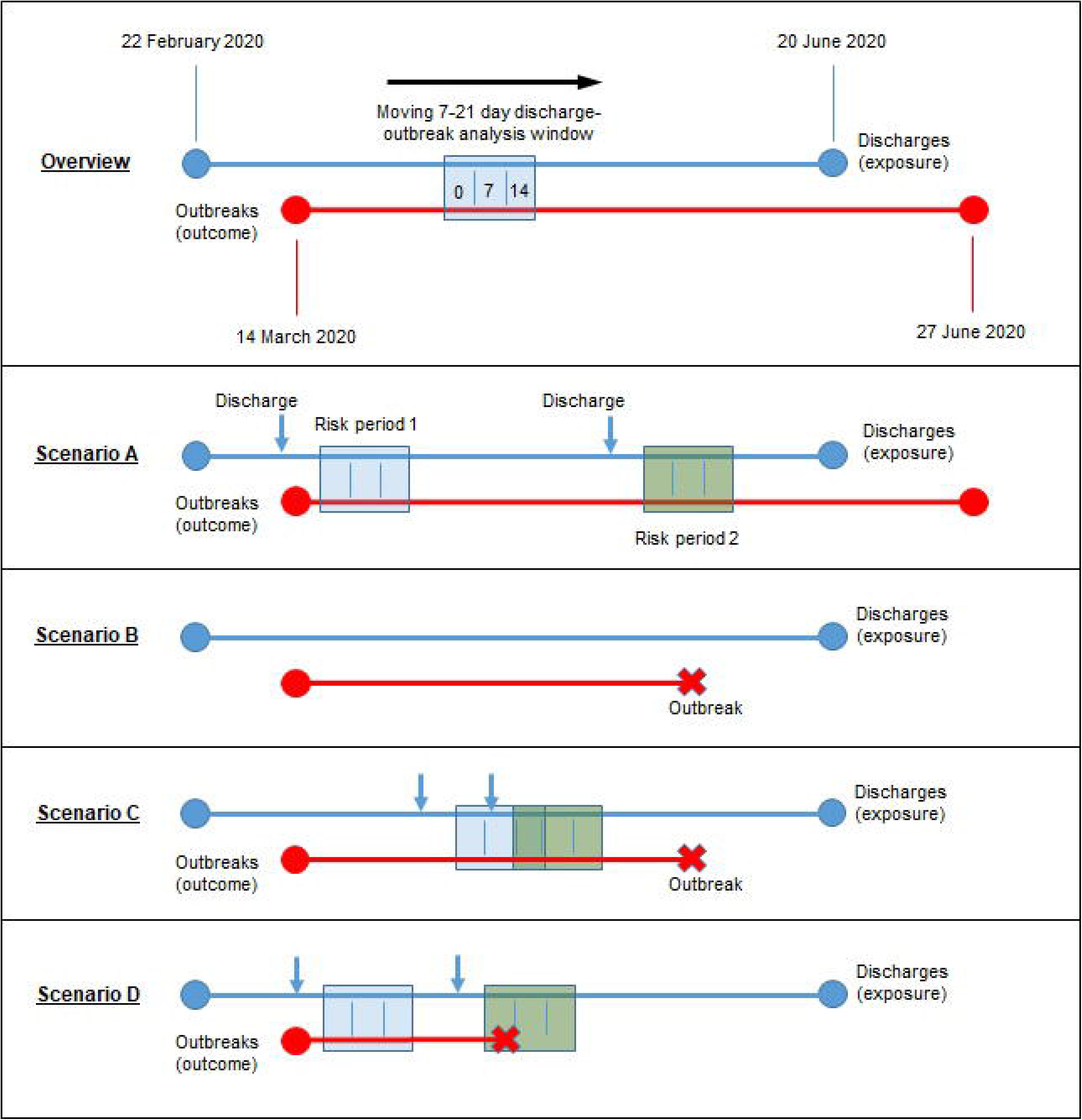
Study period analysis timeline with risk period interaction scenarios. **Scenarios:** (A) Two non-overlapping exposure periods, no outbreak; (B) No exposure to hospital discharge, outbreak occurs; (C) Two overlapping periods of exposure, outbreak occurs later when not exposed; (D) Two non-overlapping periods of exposure, outbreak occurs during the second discharge period.

## RESULTS

Of the 1,068 care homes in the analysis, an outbreak was recorded in 330 (30·9%), with a total of 1,544 recorded cases. 544 homes accepted a discharge. There were outbreaks in 245 of the 544 care homes with a discharge (45·0%). In these homes 16 experienced the outbreak prior to any discharge. There were 85 outbreaks in homes with no exposure-creating discharge (16·2%).

Of the 3,115 discharges, 1,944 were into a care home that reported an outbreak, of which 1,058 occurred prior to the outbreak and therefore created or extended a period of exposure. There were 1,171 discharges into a care home that did not report an outbreak. Dates for all 330 outbreaks and the 2,229 discharges creating or extending an exposure period between 22 February to 27 June 2020 are shown in figure 2. A summary of the characteristics and hospital discharges of the care homes in Wales is given in table 1.

**Figure 2.**
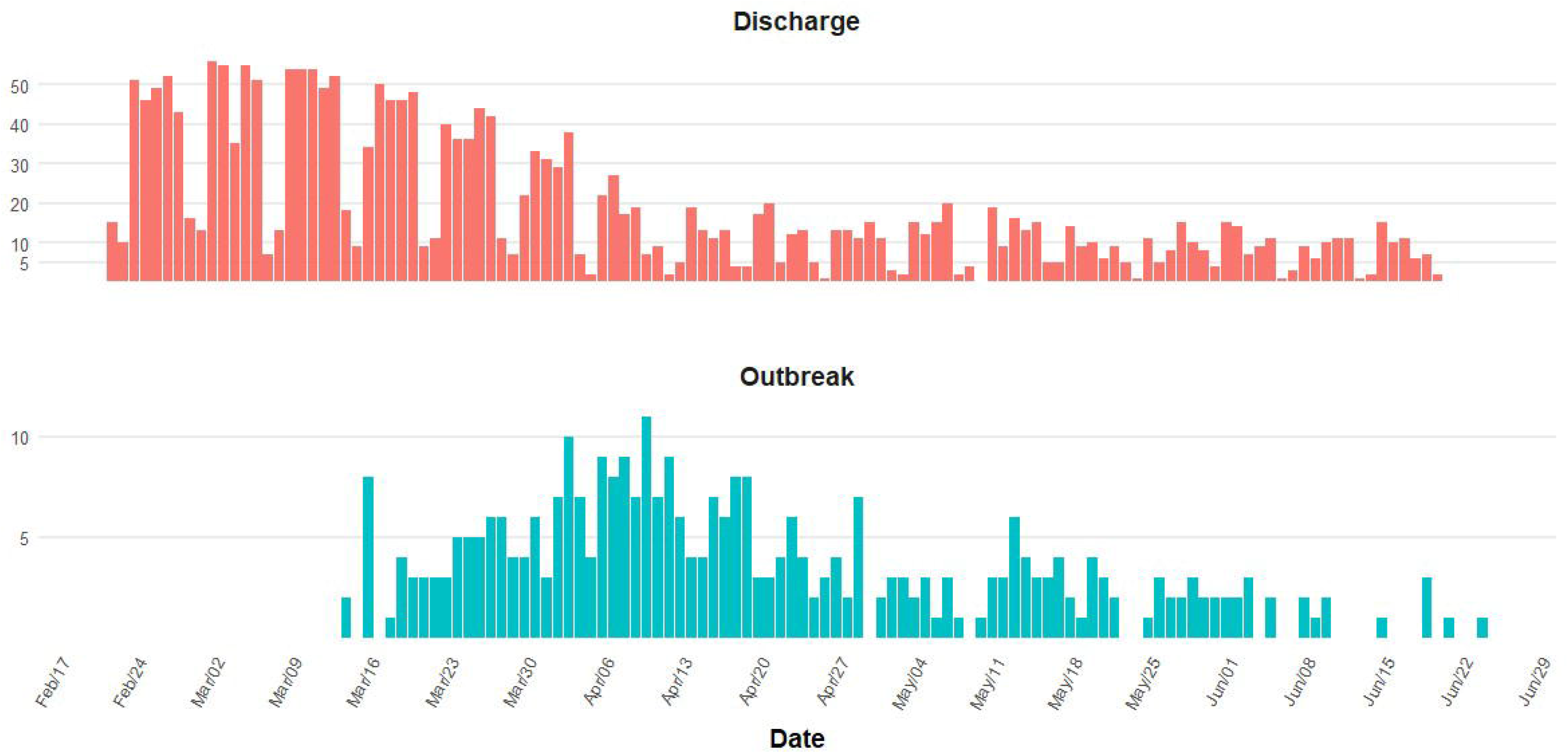
Hospital discharges creating/extending an exposure period and outbreaks (first positive SARS-CoV-2 tests in resident), care homes in Wales, 22 February to 27 June 2020.

**Table 1:** Summary statistics for hospital discharges and positive SARS-CoV-2 tests in residents of care homes in Wales, 22 February to 27 June 2020

| Health Board | Total care homes | Total capacity | Total lab confirmed results | Care homes with outbreak (n) | Care homes with outbreak (%) | Care homes with discharge (n) | Care homes with discharge (%) |
| --- | --- | --- | --- | --- | --- | --- | --- |
| Aneurin Bevan | 170 | 4,053 | 233 | 63 | 37.1 | 93 | 54.7 |
| BCU | 290 | 6,730 | 492 | 115 | 39.7 | 189 | 65.2 |
| Cardiff & Vale | 130 | 3,291 | 192 | 44 | 33.8 | 55 | 42.3 |
| CTM | 119 | 2,967 | 208 | 40 | 33.6 | 69 | 58.0 |
| Hywel Dda | 185 | 3,869 | 175 | 17 | 9.2 | 100 | 54.1 |
| Powys | 40 | 1,227 | 46 | 8 | 20.0 | 9 | 22.5 |
| Swansea Bay | 134 | 3,247 | 198 | 43 | 32.1 | 29 | 21.6 |
| **Wales Total** | **1,068** | **25,384\*** | **1,544** | **330** | **29.4** | **544** | **50.9** |

In the Cox regression, time-dependent exposure to hospital discharge in the univariable model, with no other factors, was associated with a significantly increased hazard ratio for the risk of an outbreak (2·47, 95% CI: 1·96, 3·11, see table 2). Similarly, significant univariable effects of size, dementia care, service sub type (nursing care), learning disability provision and regional health board were detected. However, in the mutually adjusted model, there was no significant association for hospital discharge, service sub type, dementia care or learning disability provision. The adjusted hazard ratio for hospital discharge was slightly raised, at 1·15, but with a 95% CI from 0·89 to 1·47 (p = 0·29). The results indicate strong confounding in the raw data by care home size, which was by far the strongest independent predictor of outbreak risk. In comparison to the reference category of small care homes with 1-9 residents, the hazard ratio for homes with 10-24 residents was 3·40 (1·99, 5·80). For homes of 25-49 the hazard ratio was 8·25 (4·93, 13·81) and for the largest category of homes (50+) it was 17·35 (9·65, 31·19). The effect of health board largely mirrored the regional size of the epidemic and therefore acted as a marker of prevalence. Proportional hazard assumptions were met in all models, as assessed by non-significant global test for time trend in residuals.

**Table 2:**
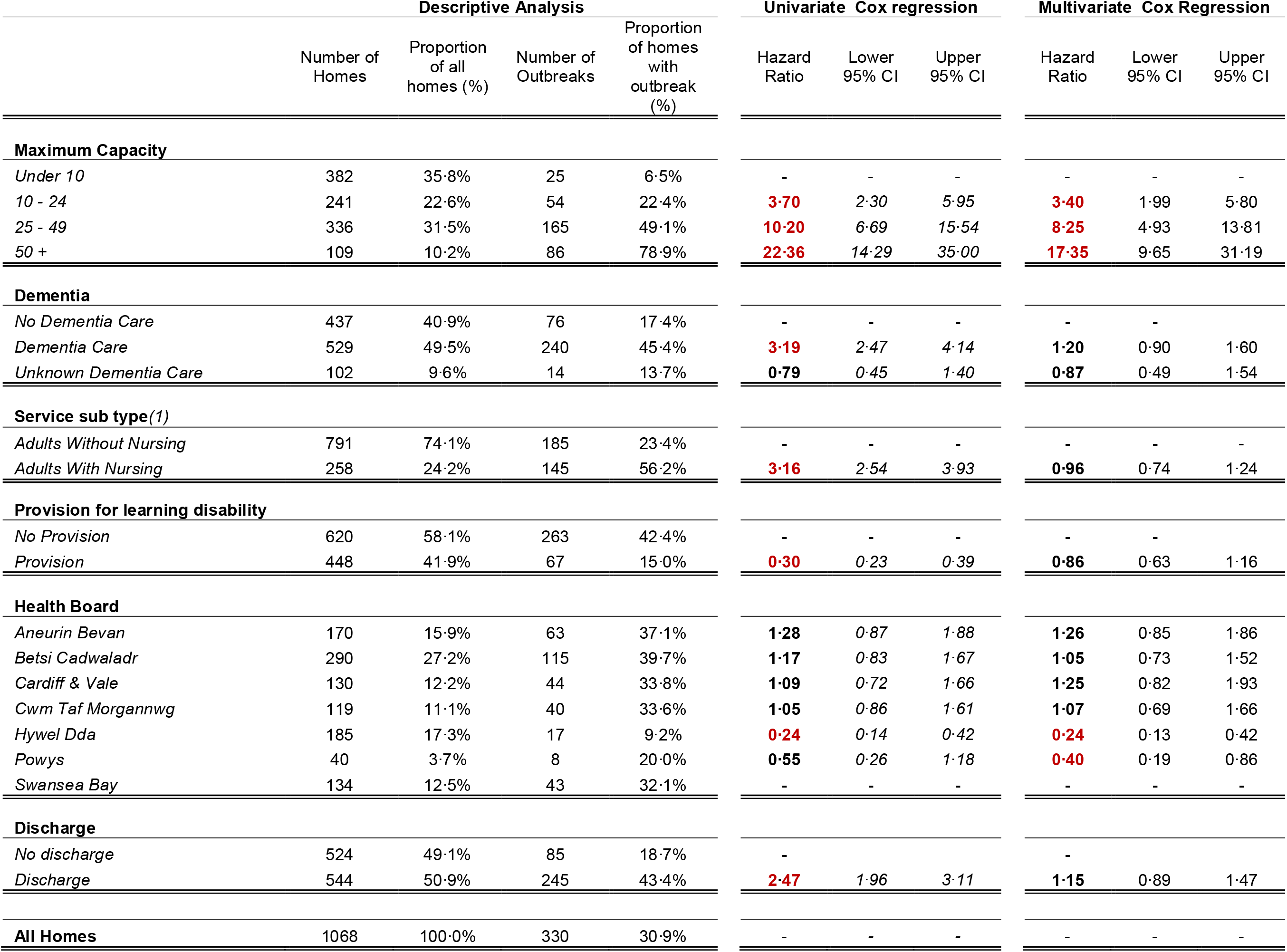
Hazard ratios for care home COVID-19 outbreaks following discharge from hospital (crude and adjusted by size and type and level of care, significant hazard ratios highlighted in red), Wales, 29 February to 27 June

The confounding effect of care home size on observed univariable effect of hospital size can clearly be seen by considering the outbreak event rate per 1000 days at risk from hospital discharge (within the window) and comparing it to the event rate when not exposed. Over all care homes, there was a recorded 6·67 outbreaks per 1000 days of exposure to hospital discharge, compared to 2·47 outbreaks per 1000 days not in the exposed window. However, after stratifying by home size there were no significant differences at any care home size category. For example, the largest (50+) care homes recorded 14·05 (95% CI 10·08, 18·22 per 1000 days when exposed to a hospital discharge, and a similar 11·69 (95% CI 8·53, 14·99) outbreaks per 1000 days when unexposed (see Table 3).

**Table 3.** – Outbreaks by care home capacity, within and outside of risk period, 22 February to 27 June 2020

| Maximum Capacity | Risk Period (7-21 days) | Days in study | Number of Outbreaks | Rate of outbreaks per 1000 days | Lower 95% Confidence interval | Upper 95% Confidence interval |
| --- | --- | --- | --- | --- | --- | --- |
| **Under 10** | Not in Risk Period | 42,830 | 23 | 0.54 | 0.34 | 0.76 |
|  | Within Risk Period | 1,148 | 2 | 1.74 | 0.21 | 5.48 |
| **10 - 24** | Not in Risk Period | 21,020 | 43 | 2.05 | 1.48 | 2.64 |
|  | Within Risk Period | 4,317 | 11 | 2.55 | 1.27 | 4.22 |
| **25 - 50** | Not in Risk Period | 19,716 | 105 | 5.33 | 4.36 | 6.26 |
|  | Within Risk Period | 8,718 | 60 | 6.88 | 5.25 | 8.53 |
| **Over 50** | Not in Risk Period | 3,850 | 45 | 11.69 | 8.53 | 14.99 |
|  | Within Risk Period | 2,919 | 41 | 14.05 | 10.08 | 18.22 |
| **All Homes** | Not in Risk Period | 87,417 | 216 | 2.47 | 2.15 | 2.76 |
|  | Within Risk Period | 17,102 | 114 | 6.67 | 5.50 | 7.79 |
| **Total** | **All** | 104,519 | 330 | 3.16 | 2.83 | 3.46 |

In our sensitivity analysis, considering a wide range of possible time-dependent exposure windows, no q values for hospital discharge reached significance at either the 5% or 10% level. The estimated overall proportion of true null hypotheses (π_0_), was 1·0. The smallest q value was 0·14, associated with an observed hazard ratio of 1 43 at a window of 10 to 31 days, and implying a minimum false discovery rate (fdr) of 14% incurred if considered significant. Very similar results were obtained using the local false discovery rate, and the estimated π_0_ remained 1·0 across all values of fdr tuning parameter λ. We note that when considering only hospital discharge and care home size in the model (omitting all other non-significant covariate) the results were almost identical. Finally, we considered the effect of the change in policy to mandate testing prior to discharge (02/05/20) by fitting the models with a factor for the two time periods. This factor was not found to be significant, and did not significantly alter hazard ratios.9

## DISCUSSION

Consistent with our study, care home size has been the only factor consistently reported as influencing the risk of outbreak, with Burton et al.^7^ describing an odds ratio for outbreak of 3·50 (95%CI 2·06 to 5·94) per 20-bed increase and Dutey-Magni et al^19^ finding an adjusted hazard ratio for individual infection of 1·59 (95%CI 2·06 to 5·94) in 45–59 bed facilities and 1·87 (95%CI 1·44 to 2·43) for 70–84 beds when compared with 20–34 bed care homes. It is possible that, because larger homes require more staff and have potentially higher levels of mixing than smaller homes, outbreaks are more likely. These homes are potentially more likely to use agency staff to fill rotas that smaller homes, who might possibly work at more than one care home and present increased opportunities to introduce infection to care homes. Homes serving residents with higher needs would be expected to have higher staff/resident ratios and be less ability to reduce their personal risk of infection through handwashing and minimising social contacts, which could be a possible reason for increased likelihood of infections in these settings. These structural and operational parameters of care homes are areas which warrant further investigation. It must be noted that although a large number of care homes and events were included in the analysis, the precision of our estimated hazard ratio for the effect of hospital discharge covers the confidence interval 0·9 to 1·5. Hence an effect within this range cannot be ruled out, and in individual cases the source of the introduction to the home could have been hospital discharge. While it is possible that few infectious cases were discharged, or they were late in infection so not excreting, it is also possible that care home staff took specific action receiving discharged patients meaning these residents were successfully isolated in the homes. In addition, the 12 potential increased risk of acquiring COVID-19 had they not been discharged to care homes should be considered. Remaining in hospital is not without risk, and there was a rationale for expediting discharges, given the expected influx of COVID-19 cases to hospitals in Wales.

### Limitations

Clearly not all discharges would have had COVID-19, so the effect of our defined risk factor would be diluted by non-risk discharges. However, the aim was to see an overall effect of the pattern and policy of discharges. It was not possible to ascertain if the case in outbreaks was the resident who had been discharged from hospital within the period of interest, and this will be the focus of further investigation. The matching of cases to discharges could be investigated to assess if a case was the primary case discharged from hospital or a secondary infection within the home. Further study will focus on understanding how many homes care home staff worked in during the study period, especially if agency staff were working across multiple homes each week. Here, we focused on the timing of the first outbreak. An analysis of the timeline of all cases is complicated by very limited information on the balance of internal and external exposure, as well as changing testing practices. Such an analysis could shed light on the time-dependent intensity of cases, and what external factors may have been contributing to that.

### Conclusions and recommendations

Larger homes were at considerably greater risk of COVID-19 outbreaks, but for the period studied, the risk was not significantly increased in the period following a hospital discharge. Further analyses should investigate the risk where discharges were confirmed or probable cases of COVID-19, and also consider additional evidence on likely chains of transmission that may become available from sources such as greater record linkage and viral genetic sequence data. Alternate sources for seeding residential care outbreaks should be investigated, including the risks to and from staff and the overlap with other community transmission. Patients who are infectious with COVID-19 or other infections can seed outbreaks into residential care and other settings, so strict policies on testing and isolation are very important to avoid outbreaks. Some of the outbreaks documented here may have been due to hospital discharges. However, overall, these discharges were not a significant factor in the spread of COVID-19 to resident care in Wales.

## Data Availability

Datasets included patient identifiable information. These were cleaned and anonymised before performing analysis on them. The anonymised data used for analysis is held on Public Health Wales servers and could be provided in principle to third parties.

## Footnotes

### Correction notice

**Contributors** CW and BM conceived the study and designed it with support from JA and MG. VM and BT extracted the hospital discharge data. CE extracted the data on care home characteristics and test results and cleaned and prepared all data for analysis. DT prepared and analysed the data with statistical testing and coding support for MG. JA wrote the first draft and edited the manuscript and all authors provided critical revisions. All authors read and approved the submitted manuscript. Care Inspectorate Wales provided information on dementia and specialist care services in Wales.

### Ethics

Ethical oversight of the project was provided by PHW R&D Division. As this work was carried out as part of the health protection response to a public health emergency in Wales, using routinely collected surveillance data, PHW R&D Division advised that NHS research ethics approval was not required. The use of named patient data in the investigation of communicable disease outbreaks and surveillance of notifiable disease is permitted under Public Health Wales’s Establishment Order. Data were held and processed under Public Health Wales’ information governance arrangements, in compliance with the Data Protection Act, Caldicott Principles and Public Health Wales guidance on the release of small numbers. No data identifying protected characteristics of an individual were released outside Public Health Wales.

### Patient consent for publication

Not required.

### Provenance and peer review

Not commissioned.

### Dissemination declaration

Public Health Wales is working closely with Welsh Government and other stakeholders including Care Inspectorate Wales in the response to COVID-19. The results of this study will be made available to all of these stakeholders through the appropriate channels.

